# Engagement and adherence trade-offs for SARS-CoV-2 contact tracing

**DOI:** 10.1101/2020.08.20.20178558

**Authors:** Tim C. D. Lucas, Emma L. Davis, Diepreye Ayabina, Anna Borlase, Thomas Crellen, Li Pi, Graham F. Medley, Lucy Yardley, Petra Klepac, Julia Gog, T. Déirdre Hollingsworth

**Affiliations:** Big Data Institute, Li Ka Shing Centre for Health Information and Discovery, University of Oxford, UK; MathSys CDT, University of Warwick, UK; Department of Infectious Disease Epidemiology, London School of Hygiene and Tropical Medicine, UK; Centre for Mathematical Modelling of Infectious Disease and Department of Global Health and Development, London School of Hygiene and Tropical Medicine, London, UK; Department for Applied Mathematics and Theoretical Physics, University of Cambridge, UK; School of Psychology, University of Southampton, Southampton, UK; School of Psychological Science, University of Bristol, UK

**Author notes:** Email address (Tim C. D. Lucas). Authors contributed equally (Tim C. D. Lucas, Emma L. Davis).

**Keywords:** COVID-19, contact tracing, branching processes, SARS-CoV-2, adherence, engagement, case isolation, quarantine

## Abstract

Contact tracing is an important tool for allowing countries to ease lock-down policies introduced to combat SARS-CoV-2. For contact tracing to be effective, those with symptoms must self-report themselves while their contacts must self-isolate when asked. However, policies such as legal enforcement of self-isolation can create trade-offs by dissuading individuals from self-reporting. We use an existing branching process model to examine which aspects of contact tracing adherence should be prioritised. We consider an inverse relationship between self-isolation adherence and self-reporting engagement, assuming that increasingly strict self-isolation policies will result in fewer individuals self-reporting to the programme. We find that policies that increase the average duration of self-isolation, or that increase the probability that people self-isolate at all, at the expense of reduced self-reporting rate, will not decrease the risk of a large outbreak and may increase the risk, depending on the strength of the trade-off. These results suggest that policies to increase self-isolation adherence should be implemented carefully. Policies that increase self-isolation adherence at the cost of self-reporting rates should be avoided.

## 1. Background

Since the first cases of SARS-CoV-2 in China in late 2019^1^ the virus has spread globally, resulting in over 600,000 confirmed deaths by August 2020^2^. Lockdown in the UK began in March 2020^3^ and reduced *R*_0_ below 1 while also triggering unprecedented reductions in economic activity^4^. As lockdown restrictions are relaxed, both in the UK and in other countries, other methods for keeping *R*_0_ below one are needed. Large-scale contact tracing is one of the potential methods for keeping virus spread under control^5,6,7^.

During the current SARS-CoV-2 outbreak, contact tracing has been used to great effect in a number of countries including Vietnam and South Korea^8,9^. Two broad classes of contact tracing include manual tracing and digital tracing using a smartphone app^10^. Manual contract tracing is the only system currently running in the UK though it is expected that a contact tracing app will be launched soon. In manual contact tracing, trained public health staff ask a case for the names and contact details of people they have recently been in close proximity with, as well as asking for information on which public areas the infected person has visited. The tracers will then identify as many contacts as possible and ask them to self-isolate for a period. Adherence to the contact tracing system is an important determinant of its efficacy^11,6,10^.

Adherence applies to a number of different aspects of contact tracing^12^. Untraced individuals with symptoms must report themselves to the contact tracing system. They then must give identifying information about the people they have been in close proximity with. Then, both the index case, and the traced contacts, must self-isolate for a period^13,14^. If the contact tracing system uses home swab tests, the swabs must be taken carefully^15,16^. Adherence to each of these steps will be imperfect.

Although there are many unobserved variables involved, we can start to examine some of these adherence rates using public statistics from the UK tracing system^17^. For example, of the 6,923 people who were referred to the contact tracing system between the 11th and 17th of June, 70% were reached. However, these 6,923 cases certainly do not represent 100% of the new cases in the country that week. Of the 6,923, 74% gave details of at least one contact though it is not possible to tell how many of the remaining 26% actually had no contacts. 82% of close contacts were reached and asked to self-isolate.

However, these adherence rates are not fixed parameters and can be influenced by policy. For example, economic support for those missing work^14,18^, daily phonecalls to monitor adherence^19^ or legal ramifications for breaking self-isolation, such as those implemented in Singapore and Taiwan^19^, might be expected to increase self-isolation rates^18^. In particular, this work was originally undertaken in response to a question from policy makers asking whether legally mandating self-isolation for close-contacts would reduce transmission rates. Furthermore there are likely to be trade-offs and dependencies between parameters. In particular, contact tracing relies on self reporting of symptoms in order to initially identify a chain of transmission but introducing penalties for non-compliance to self-isolation might be expected to decrease the proportion of people that report themselves to the system in the first instance. In general there are few direct, individual benefits to self reporting oneself to a contact tracing system; instead the benefits are communal and the drivers for self reporting are likely to be altruism or social norms^20,21^. However, there are direct costs both to the individual that self reports and to their close contacts. Self-isolation is mentally difficult^22^ and will come with economic costs for many^20,23,24,14,25^. Legally enforcing self-isolation exacerbates these costs.

The exact form that these trade-offs would take are difficult to know. Adherence to self-isolation requirements might largely be binary with people complying for the full 14 days (as requested in the UK) or not adhering at all. In this case, legal enforcement would be expected to increase the proportion of people that self-isolate. Alternatively, it is possible that self-isolation adherence is more continuous with people adhering for a few days instead of the full 14 days. Similarly, legal enforcement might be expected to increase the duration of isolation. Finally, if swab tests are being self-administered, people might be less careful or less willing to endure discomfort if the consequences of a positive test are more severe (though this might change as saliva tests are produced^26,27^). While it is difficult to know the functional effects of different levels of compliance, it is even more difficult to quantify the strengths of the trade-offs. Legal enforcement might have a weak effect on improving self-isolation adherence^22^ but a strong deterrent effect on self-reporting. Alternatively, perhaps legal mandation has a strong effect on self-isolation adherence without being a strong deterrent to self-reporting rates Furthermore, the shapes of these trade-offs are likely to differ in different countries and social groups based on culture, trust in the government and other factors. Careful quantitative and qualitative studies will need to be conducted to quantify these effects.

Here we use a previously published branching process model^11,6^ to examine the effects of these trade-offs on the risk of a large outbreak of SARS-CoV-2. We examine trade-offs between self-isolation duration and self-isolation probability with self-reporting rates, contact information reporting probabilities and sensitivity of home swab tests. It is important to note however that we do not consider the societal costs^28^ of legal enforcement of self-isolation; we aim to quantify the benefits of these policies without considering the costs noting that the costs are not easy to directly compare to the benefits.

## 2. Methods

In this paper we extend a previous model of SARS-CoV-2 transmission^11^. An overview of the model is given in Figure S1 while parameter values and references are given in Table 1. At the individual-level, the number of potential secondary contacts are modelled by a negative binomial distribution while the exposure times of these new infections are modelled as a gamma distribution. Self-isolating individuals are assumed to be unable to transmit the disease (assuming isolation within households) and therefore potential secondary cases are avoided if the gamma-distributed exposure time occurs during self-isolation of the primary case. The timing of self-isolation depends on whether the case was traced as a potential contact or not and a number of factors affecting adherence as described in detail below. The model proceeds as a branching process with each simulation being seeded with twenty untraced, infected individuals.

**Table 1:**
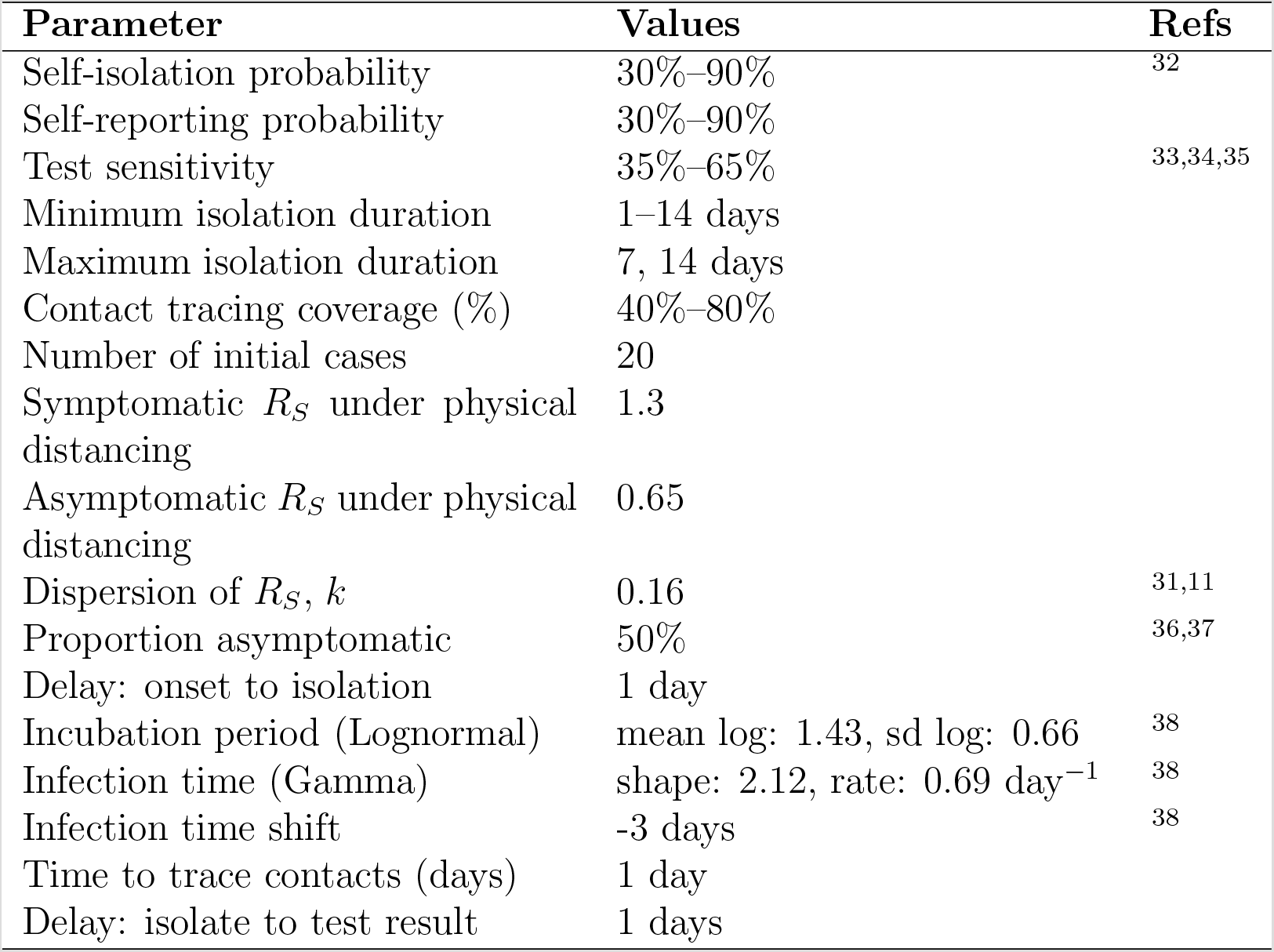
Model parameters values/ranges. Parameters taken from the literature are fixed and for other parameters a range of values are explored.

### 2.1. Secondary case distribution

The heterogeneity in the number of potential secondary cases caused by an individual is modelled as a negative binomial distribution. For symptomatic cases we use a mean value of 1.3 secondary cases while asymptomatic cases are given a 50% lower infection rate. This relates to a scenario where strong social distancing and good hygiene is still being observed. Earlier work^6,7^ and preliminary analyses indicated that contact tracing is unable keep the risk of an outbreak low without being paired with social distancing so this is the scenario we focus on. Estimates for the dispersion parameter, *k*, for SARS-COV-2 range from *k* = 0.1 (0.05–0.2) for pre-lockdown UK^29^ to *k* = 0.25 (0.13–0.88) for Tianjin, China during lockdown measures^30^. Given this range we have kept the parameter as used in^11,31^ setting *k* = 0.16. This value of *k* yield a strongly skewed distribution with most individuals causing zero potential secondary cases.

### 2.2. Infection profile

Individuals are labelled as symptomatic or asymptomatic with a probability of 50%^36,37^. The onset time of symptoms is modelled as a lognormal distribution with mean 1.43 days and sd of 0.66^39^. All individuals, whether symptomatic or asymptomatic are given a symptom onset time as the exposure time of secondary cases is calculated relative to this time. The exposure time for each new potential case is drawn from a gamma distribution with shape parameter of 17.77 and a rate of 1.39 day^*−*1^. This distribution is centred three days before the onset of transmission^38^. The sampled exposure time is compared to the infector’s exposure time and resampled if it occurs before the infector was infected. If the exposure time of a potential secondary case occurs during the primary case’s self-isolation, the infection event does not occur and the Ppotential secondary case does not become a case. These distributions are shown in Figure S2.

**Figure 1:**
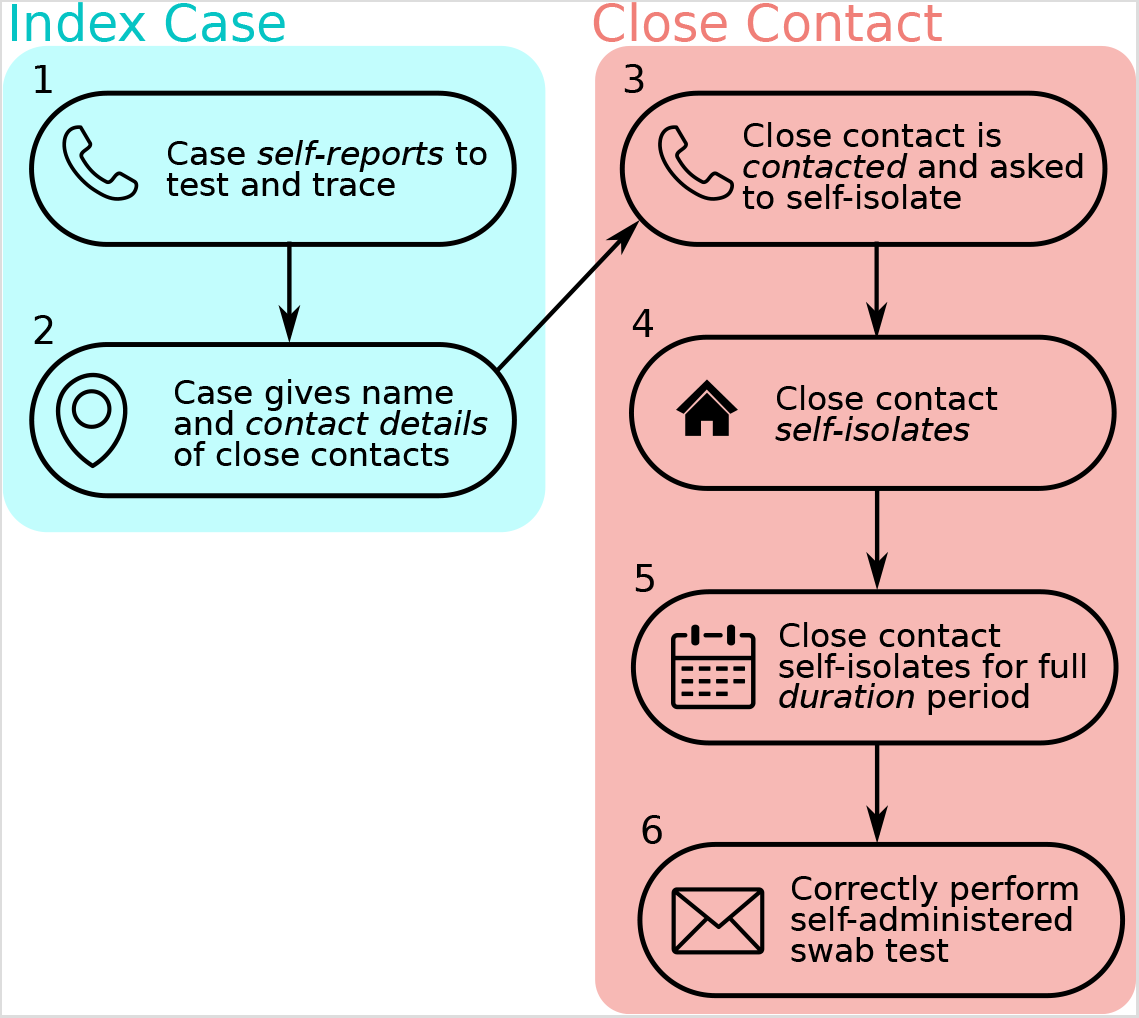
Overview of adherence in test and trace. An untraced individual must self-report and give the name and details of close contacts. The contact tracing team must then manage to contact the close contacts. The close contacts must self-isolate when asked and remain in self-isolation for the full isolation period (14 days in the UK). In some systems, the isolated individual is given a self-administered swab test which must be administered correctly. There is imperfect adherence or performance at each of these stages. In this paper we focus on trade-offs between self-report rate (stage 1) and self-isolation adherence (stages 4 and 5). We combine stages 2 and 3 into one parameter, which we call control effectiveness.

### 2.3. Contact tracing

The first stage in the contact tracing system is an untraced, symptomatic individual self-reporting themselves. The control effectiveness, i.e. the proportion of an individual’s epidemiological contacts that are recalled, divulged and successfully contacted by the contact tracing team, is varied between 40%–80%. If contact tracing is successful, the traced individual is asked to self-isolate. We assume it takes one day to contact a contact. If a traced contact subsequently shows symptoms or returns a positive test the next round of contact tracing is initiated. That is, the contacts of the traced contact are then traced.

### 2.4. Testing

As a baseline we assume that tests have a sensitivity of 65% and that it takes one day for results to be returned. This reflects the sensitivity of tests observed in the community^34,33^. Given a positive test result contact tracing for the tested individual is initiated. A negative test allows the tested individual to be immediately released from quarantine. Any contacts of a negative-testing case that were successfully identified prior to receiving the test result are still isolated and tested. In a branching process model only infected individuals are modelled. Therefore we do not track the number of uninfected people that are unnecessarily asked to quarantine. Test specificity affects the number of uninfected people asked to quarantine but does not directly affect the spread of the disease and therefore we do not define a test specificity. In this study we are concerned with quantifying the benefits of contact tracing and do not attempt to weigh the epidemiological benefits against the sociological costs.

### 2.5. Adherence trade-offs

We consider three main trade-offs. As we do not have good data to define the shapes of these trade-offs we run simulations for all combinations of parameters.

First, we assume that without policies to encourage self-isolation most people attempt some self-isolation but the lack of adherence is with respect to the duration of self-isolation that decreases. We keep the probability of self-isolation constant at 70%. We assume that each person that does self-isolate isolates for an amount of time taken from a uniform distribution between a minimum and maximum value. For the maximum values we use either the full 14 days currently recommended in the UK or a shorter seven day maximum isolation. We vary the minimum duration of self-isolation from 1 day to being equal to the maximum duration.

Second, we examine the trade-off between self-report probability and self-isolation probability. We expect that policies that increase self-isolation probability will reduce self-report probability. We use values of self-isolation from 10% – 70% in increments of 20% and examine all combinations with self-report probabilities from 10% – 70% also in increments of 20%. The upper bound for self-isolation here is certainly above the rate of self-isolation currently being achieved in the UK. However, it is below the target rate for other national contact tracing programmes^32^. Furthermore, the very strict restrictions applied to travellers entering countries such as Singapore could also be considered an upper bound on feasible policies. Many of the policies used in these areas, such as enforced isolation in government run hotels, GPS ankle bracelets, and daily video calls, would be considered draconion if applied to the population at large but could be reasonably expected to produce self-isolation rates of 90%. In contrast to the first trade-off, we assume that everyone that does self-isolate does so for the full maximum value of either 7 or 14 days.

Finally, we assume that policies that increase self-isolation probability will decrease test sensitivity. This scenario applies to the case of home administered tests. With strong incentives to test negative, people will be less likely to perform swabs correctly. We therefore examine a range of test sensitivities from a baseline of 65% down to 35% in increments of 10%.

### 2.6. Simulation process

Results presented are the combined output of 15,000 simulations for each parameter combination, or scenario, considered. We define a simulation as leading to a large outbreak if it has more than 2,000 cumulative cases or if there are still infected cases after 300 days. The threshold of 2,000 cases was chosen by running simulations with a maximum of 5,000 cases and noting that of the simulated epidemics that went extinct, 99% of extinction events occurred before reaching 2,000 cases. Nearly all simulations either went extinct or reached 2,000 cases with very few simulations lasting longer than 300 days. These simulations were then used to calculate the probability of a large outbreak given a certain set of conditions. 95% Clopper-Pearson exact confidence intervals were also calculated. The model was written in R and the code and testing suite^40^ is publicly available on GitHub (https://github.com/timcdlucas/ringbp/tree/adherence_tradeoff_runs).

## 3. Results

### 3.1 Trade-off between self-isolation duration against self-report probability

We find that increasing the duration of self-isolation increases the risk of a large outbreak in the presence of reductions in self-reporting rates. The probability of a large outbreak for all combinations of self-isolation duration and self-report rates are shown in Figure 2. If we assume that we are currently in the top left panel (high self report rates but isolation taken uniformly between 1 and 14 days), policies that move us down and right generally increase the risk of a large outbreak. For example, if we consider a control effectiveness of 60%, with a self-isolation duration of between 1 and 14 days and a self-report rate of 70% the risk of a large outbreak is 1%. If we increase the self-isolation duration to always be 14 days but reduce the self-report rate to 10%, the probability of a large outbreak increases from 1% to 6%. If the trade-off is very weak, such that increasing self-isolation duration to always be 14 days only decreases self-report rates to 50%, we see no change in the probability of an outbreak.

**Figure 2:**
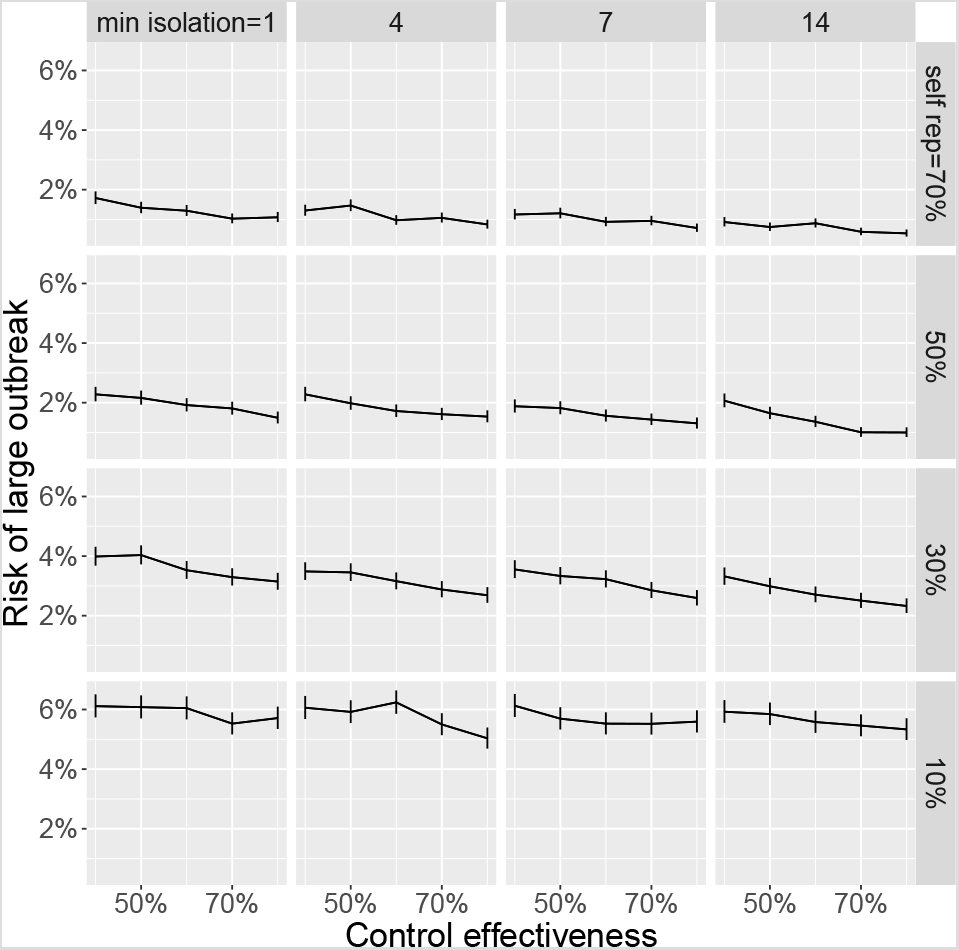
Trade-off between self isolation time (columns) and self report rate (rows) with error bars denoting 95% confidence intervals. Individuals self isolate for a randomly selected duration between min isolation and 14 days. Untraced, symptomatic individuals self-report with a probability that varies across the rows. The proportion of close contacts that are divulged and asked to self-isolate varies across the x-axis of each subplot. The y-axis shows the risk of a large outbreak (greater than 2,000 cases) over 15,000 simulations. The probability that an individual self-isolates at all is fixed at 70%. If we assume we are currently near the top left we expect that introducing legal ramifications for breaking self isolation to move us down and right. This generally increases risk.

If we assume a more pessimistic starting scenario of a self-isolation duration of between 1 and 14 days and self-reporting rates of 10% and given a control effectiveness of 60% we have a 6% risk of a large outbreak. We find that increasing self-report rates gives a larger reduction in risk. Increasing self-report rates from 10% to 70% reduces risk from 6% to 1%. In contrast, increasing the duration of isolation to always being 14 days does not change the risk of a large outbreak. We find that reducing the maximum isolation duration from 14 days to 7 days consistently increases the risk of a large outbreak (Figure S3–S5).

### 3.2 Trade-off between self-isolation probability against self-report probability

We find that increasing self-isolation probability while decreasing self report probability does not strongly alter the probability of a large outbreak. The probability of a large outbreak for all combinations of self-isolation rates and self-report rates are shown in Figure 3. If we assume that we are currently in the top left panel (high self report rates but low self-isolation rates), policies that increase self-isolation rates but decrease self-report rates would move us down and right. However, whether this decreases the risk of an outbreak depends on the strength of the trade-off. For example, if we consider a control effectiveness of 60%, with a self-isolation rate of 10% and a self-report rate of 70% the risk of a large outbreak is 6%. If we increase the self-isolation rate to 70% and equivalently reduce the self-report rate to 10%, the probability of a large outbreak is still 6%. If the trade-off is weak, such that increasing self-isolation from 10% to 70% only incurs a reduction in self-report rate to 50%, the reduction in risk of a large outbreak is substantial, reducing from 6% to 1.5%. However, if the trade-off is strong, such that increasing self-isolation from 10% to 30% causes a reduction in self reporting rate from 70% to 10%, the risk of an outbreak instead marginally increases from 6% to 7%.

**Figure 3:**
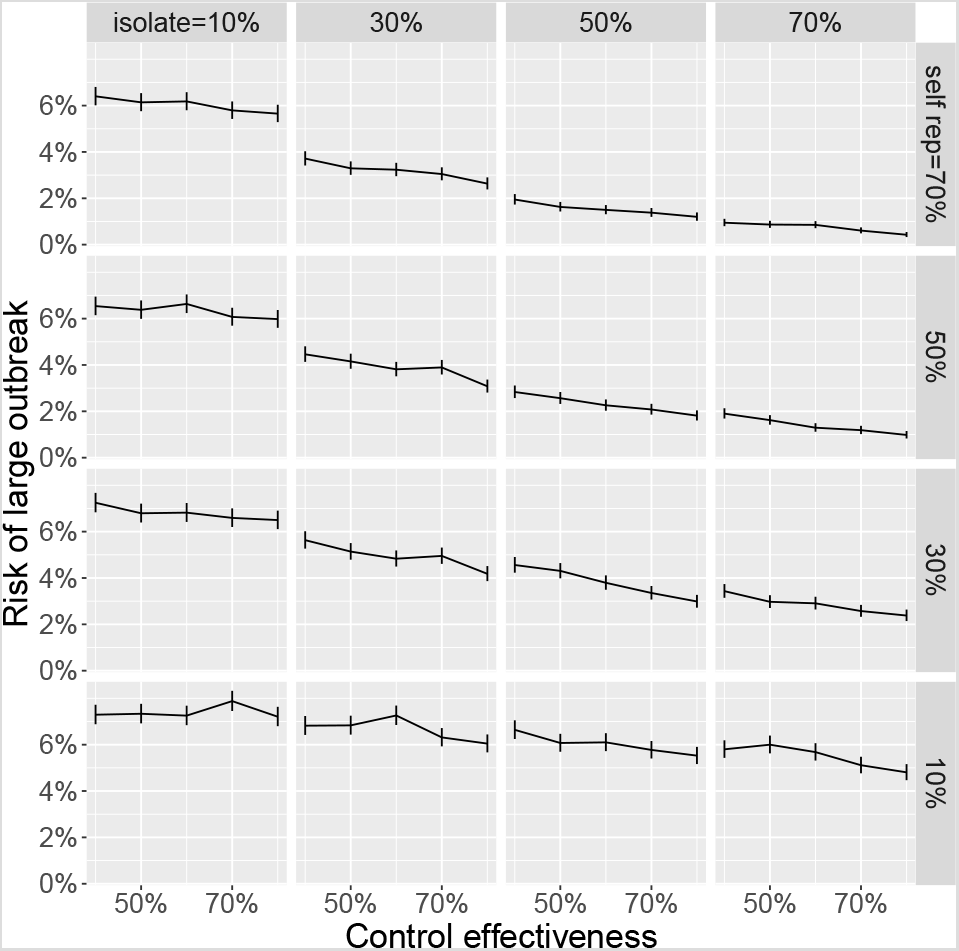
Trade-off between self-isolation probability (columns) and self-report probability (rows) with error bars denoting 95% confidence intervals. The y-axis shows the risk of a large outbreak (greater than 2,000 cases) over 15,000 simulations. If we assume we are currently near the top left we expect that introducing legal ramifications for breaking self isolation to move us down and right. Whether this decreases risk depends on the strength of the trade-off. If the trade-off is weak, such that as we move from the top left to isolation probability of 70% and self report probability of 50%, risk is reduced. In contrast, if increasing isolation probability from 10% to 30% incurs a drop in self reporting from 70% to 10%, risk does not change.

We could instead assume a more pessimistic starting scenario of self-isolation rates of 10% and self-reporting rates of 10%. Given a control effectiveness of 60% we have a 7% risk of a large outbreak. However, from this scenario we can consider whether it is better to increase self-isolation or to increase self-reporting. Increasing self isolation probability to 70% reduces risk to 6% and increasing self-report probability to 70% also reduces risk to 6%. Increasing both to 30% reduces risk to 5%. Overall, these two parameters are relatively evenly balanced. isolate = 10%

### 3.3. Trade-off between self-isolation duration against test sensitivity

To model a decrease in careful administration of home swab tests, we vary the test sensitivity and isolation adherence. We find that increasing self-isolation rates decreases the risk of a large outbreak even if this occurs in combination with reductions in test sensitivity. The probability of a large outbreak for all combinations of self-isolation rate and test sensitivity are shown in Figure 4. If we assume that we are currently in the top left panel (relatively high test sensitivity but low self-isolation rates), policies that increase self-isolation rates but decrease test sensitivity would move us down and right and this in general yields reduced risks of a large outbreak. For example, if we consider a control effectiveness of 60%, with a self-isolation rate of 10% and a test sensitivity of 65% the risk of a large outbreak is 6%. If we increase the self-isolation rate to 70% while reducing the test sensitivity to 35%, the probability of a large outbreak reduces from 6% to 3%.

**Figure 4:**
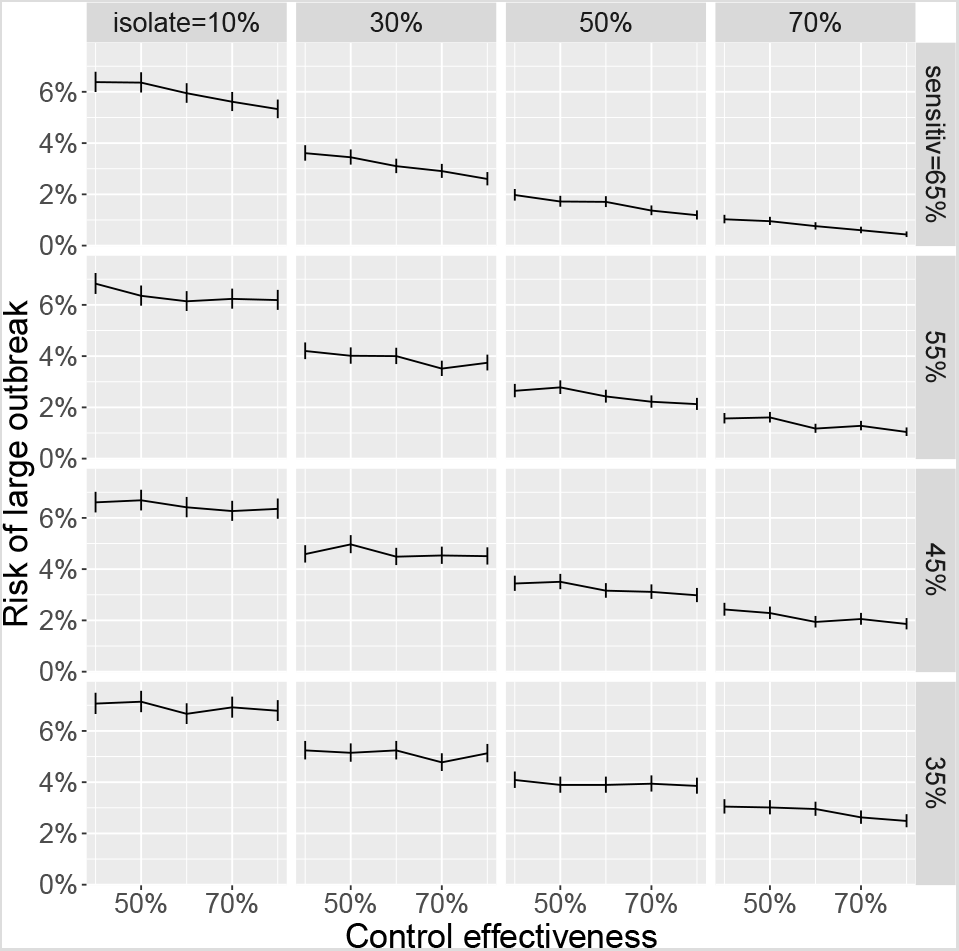
Trade-off between self isolation probability (columns) and test sensitivity (rows) with error bars denoting 95% confidence intervals. Untraced, symptomatic individuals self-report with a probability that varies across the rows. The proportion of close contacts that are divulged and asked to self-isolate varies across the x-axis of each subplot. If we assume we are currently near the top left, introducing legal ramifications for breaking self isolation might move us down and right. This generally decreases risk unless the trade off is very strong such that a small increase in isolation probability incurs a large decrease in test sensitivity.

## 4. Discussion

Overall we have found that policies that increase self-isolation rates at the expense of self-report rates are unlikely to improve the effectiveness of contact tracing systems. If the primary trade-off is between the duration of self-isolation and the probability of self-reporting, we find that policies that increase self-isolation and reduce self-report rates will cause either an increase or no change in the probability of a large outbreak depending on the strength of the trade-off. When the primary trade-off was instead between the probability of self-isolation and the rate of self report policies that increase self-isolation rates and reduce self-report rates can increase or marginally decrease the probability of a large outbreak depending on the strength of the trade-off. Overall this implies that policies such as fines, and police enforcement of self-isolation will have either little benefit or a negative effect. Broadly, policies that improve self-report rates, even at the expense of self-isolation rates should be used. This might include publicity that encourages people to self-report by reminding them that there are no legal consequences to them or their contacts for doing so.

Policies that improve self-report rates or self-isolation rates without an associated trade-off will also improve contact tracing efficacy. For example, economic support and employment protection for individuals that self-isolate would be expected to improve self-isolation rates^14,18,25^ without decreasing self-report rates. Similarly, efforts to communicate the reasons why people should self-report and self-isolate may improve both of these rates simultaneously^18,25^.

One of the core assumptions to this work is that legal consequences for breaking self-isolation would improve self-isolation rates. However, the evidence for this is not strong and there is evidence that feelings of shame do not promote adherence^21,25^. In contrast there is good evidence that other factors such as income and boredom^41^ do affect self-isolation rates. How effectively legal consequences for breaking self-isolation can increase self-isolation rates is a complex question that will depend on cultural norms, perceived enforcability, and the strength of economic and psychological consequences for self-isolation. An important consequence of this is that self-isolation rates and the effectiveness of policies aimed to improve these rates will be strongly correlated such that individuals that are most likely to infect each other are also likely to have similar self-isolation rates. This is not included in our model but has the potential to strongly reduce contact tracing efficacy in certain groups and locations.

With regards to test sensitivity, our results are relevant only to self-administered swab-tests. Swab-tests may be replaced with reliable paper-based tests. Given that we found that optimising self-isolation rates over test-sensitivity minimises risk, other considerations such as test timing and access are probably more important. Furthermore, currently in the UK, traced contacts are not allowed out of quarantine after a negative test so the system is more robust to low test sensitivity than in our simulations.

Here we have focused solely on the probability of a large outbreak as a consequence of policy change. However, there are other costs and benefits to changing values of self-report rates and self-isolation rates. High self-report rates not only improves contact tracing efficacy directly, it also creates a more effective system for measuring the incidence of SARS-CoV-2 in the community. This gives better early warning for when an outbreak is beginning in an area or group and allows for health care resources to be deployed more efficiently. In contrast, self-isolation comes with many economic and social costs both for the individual and the community. Avoiding strict penalties for breaking self-isolation allows those most affected by these costs to self-isolate less and may increase buy-in to the system as a whole. Furthermore, enforcement of self-isolation policies are an infringement on a basic liberty. While we have not tried to compare these costs to the epidemiological benefits, they must always be taken into account when implementing policy.

## Data Availability

All simulation code is available on github.
No data other than simulated data was used.

https://github.com/timcdlucas/ringbp/tree/adherence_tradeoff_runs

## 5. CRediT contribution statement

Conceptualisation: All authors

Formal Analysis: ELD, TCDL

Funding acquisition: TDH

Investigation: ELD, TCDL, AB, LP, DA, TC, LY

Methodology: ELD, TCDL

Software: ELD, TCDL

Visualization: ELD, TCDL, LY

Writing – original draft: ELD, TCDL

Writing – review & editing: All authors

## 7. Acknowledgments & funding sources

ELD, TCDL, AB, DA, LP, GFM & TDH gratefully acknowledge funding of the NTD Modelling Consortium by the Bill & Melinda Gates Foundation (BMGF) (grant number OPP1184344). The following funding sources are acknowledged as providing funding for the named authors. This research was partly funded by the Bill & Melinda Gates Foundation (NTD Modelling Consortium OPP1184344: GFM). This project has received funding from the European Union’s Horizon 2020 research and innovation programme – project EpiPose (101003688: PK). Royal Society (RP/EA/180004: PK). Wellcome Trust (210758/Z/18/Z: JH, SA). Views, opinions, assumptions or any other information set out in this article should not be attributed to BMGF or any person connected with them. TC is funded by a Sir Henry Wellcome Fellowship from the Wellcome Trust (reference 215919/Z/19/Z). Lucy Yardley is an NIHR Senior Investigator and her research programme is partly supported by NIHR Applied Research Collaboration (ARC)-West, NIHR Health Protection Research Unit (HPRU) for Behavioural Science and Evaluation, and the NIHR Southampton Biomedical Research Centre (BRC) All funders had no role in the study design, collection, analysis, interpretation of data, writing of the report, or decision to submit the manuscript for publication. This work was undertaken as a contribution to the Rapid Assistance in Modelling the Pandemic (RAMP) initiative, coordinated by the Royal Society.

## 8. Supplementary material

**Figure S1:**
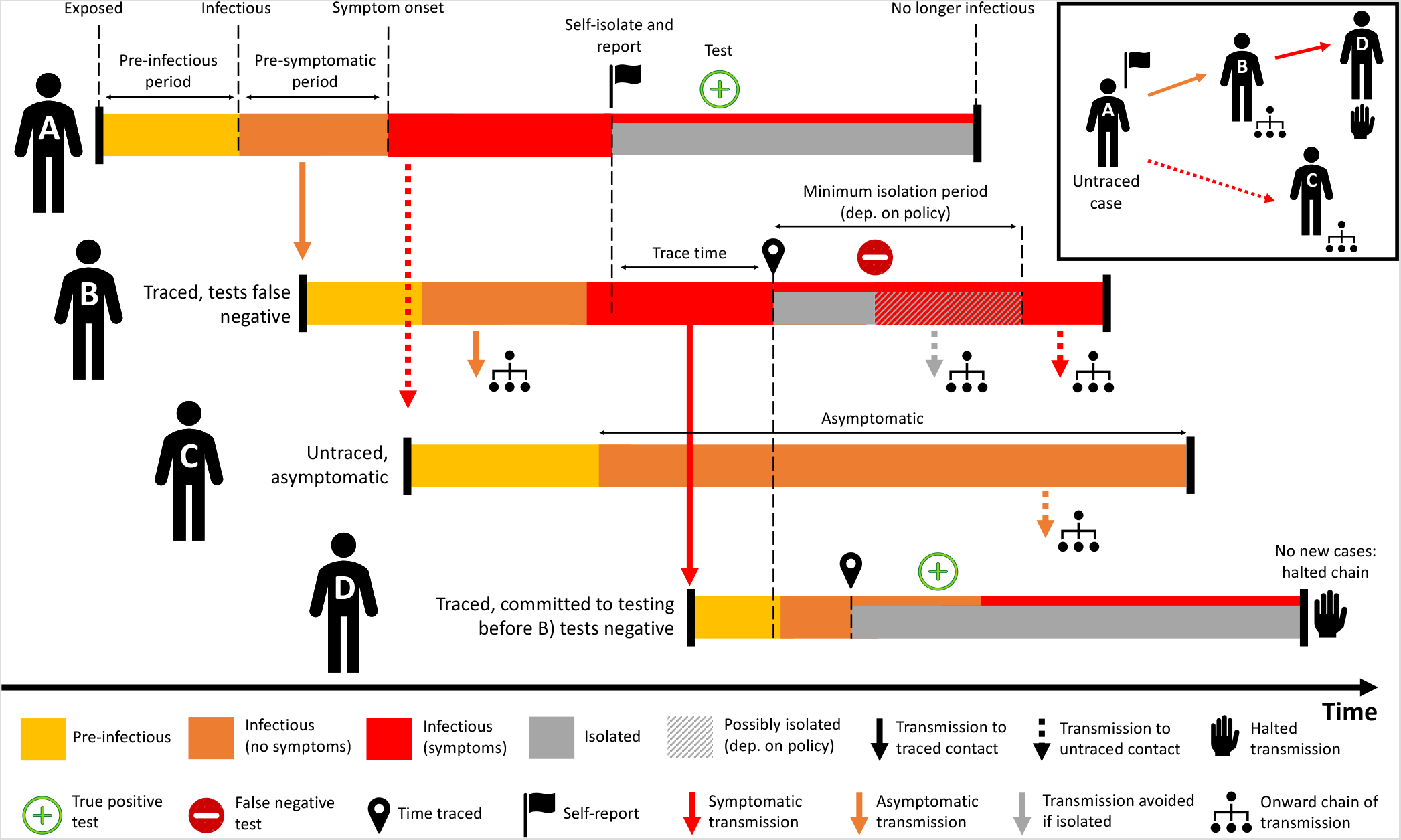
Overview of the contact tracing process implemented in our model. **Person A** isolates and self-reports to the contact tracing programme with some delay after symptom onset, by which time they have infected Persons B and C. When Person A self-reports contact tracing is initiated. They are then tested with positive result and remain isolated for their infectious period. **Person B** was infected by A prior to their symptom onset and is detected by tracing after some delay, after infecting Person D. After isolating they are tested, with a false negative result. This leads to B either a) stopping isolation immediately or b) finishing a minimum 7 day isolation period. Both may allow new onward transmission. **Person C** was infected by A but not traced as a contact. Person C does not develop symptoms but is infectious, leading to missed transmission. **Person D** was traced and tested before the false negative test was returned for Person B. The test for D returns positive, meaning that D remains isolated, halting this chain of transmission.

**Figure S2:**
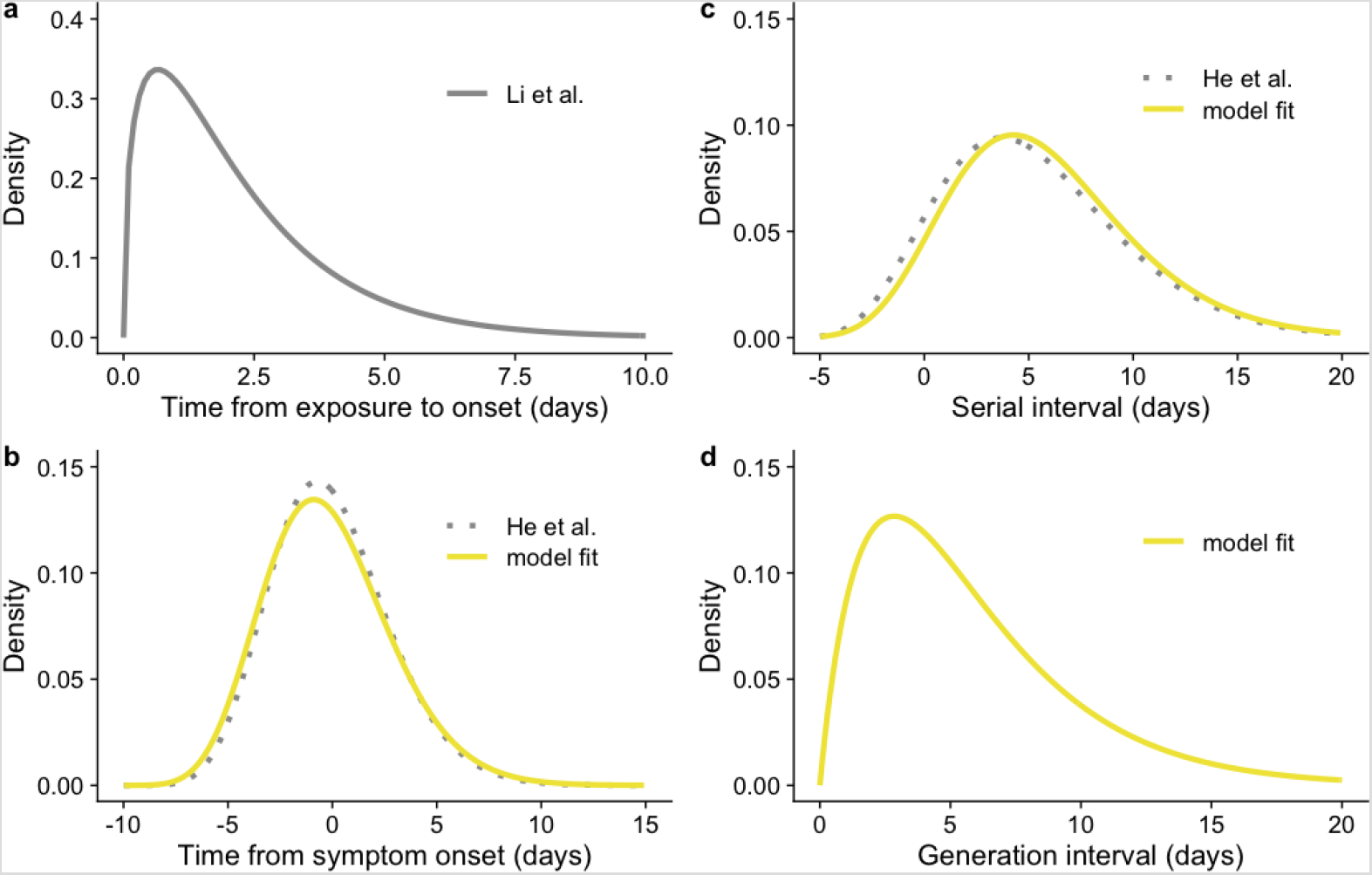
Distributions for a) incubation period (exposure time to symptom onset) from Li et al.^39^; b) transmission profile relative to symptom onset, fitted to data and compared to He et al.^38^; c) serial interval, fitted and compared to He et al.^38^; and d) generation interval, combined distribution from a) and b) with re-sampling to prevent negative serial intervals, as described in the main text.

**Figure S3:**
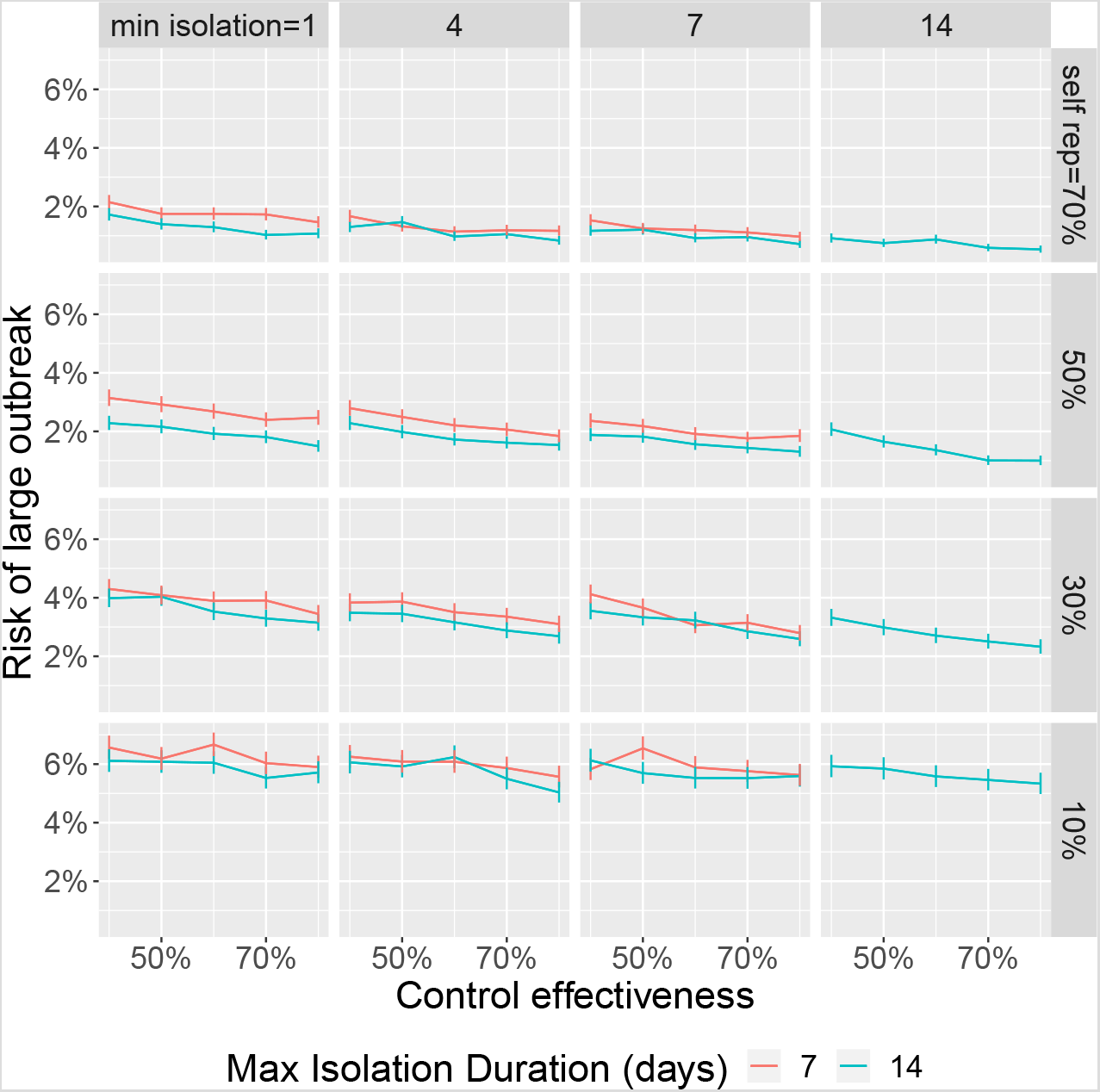
Trade-off between self isolation time (columns) and self report rate (rows) with error bars denoting 95% confidence intervals. Individuals self isolate for a randomly selected duration between min isolation4 and 14 days. Untraced, symptomatic individuals self-report with a probability that varies across the rows. The proportion of close contacts that are divulged and asked to self-isolate varies across the x-axis of each subplot. Self isolation probability is fixed at 70%. If we assume we are currently near the top left we expect that introducing legal ramifications for breaking self isolation to move us down and right. This generally increases risk.

**Figure S4:**
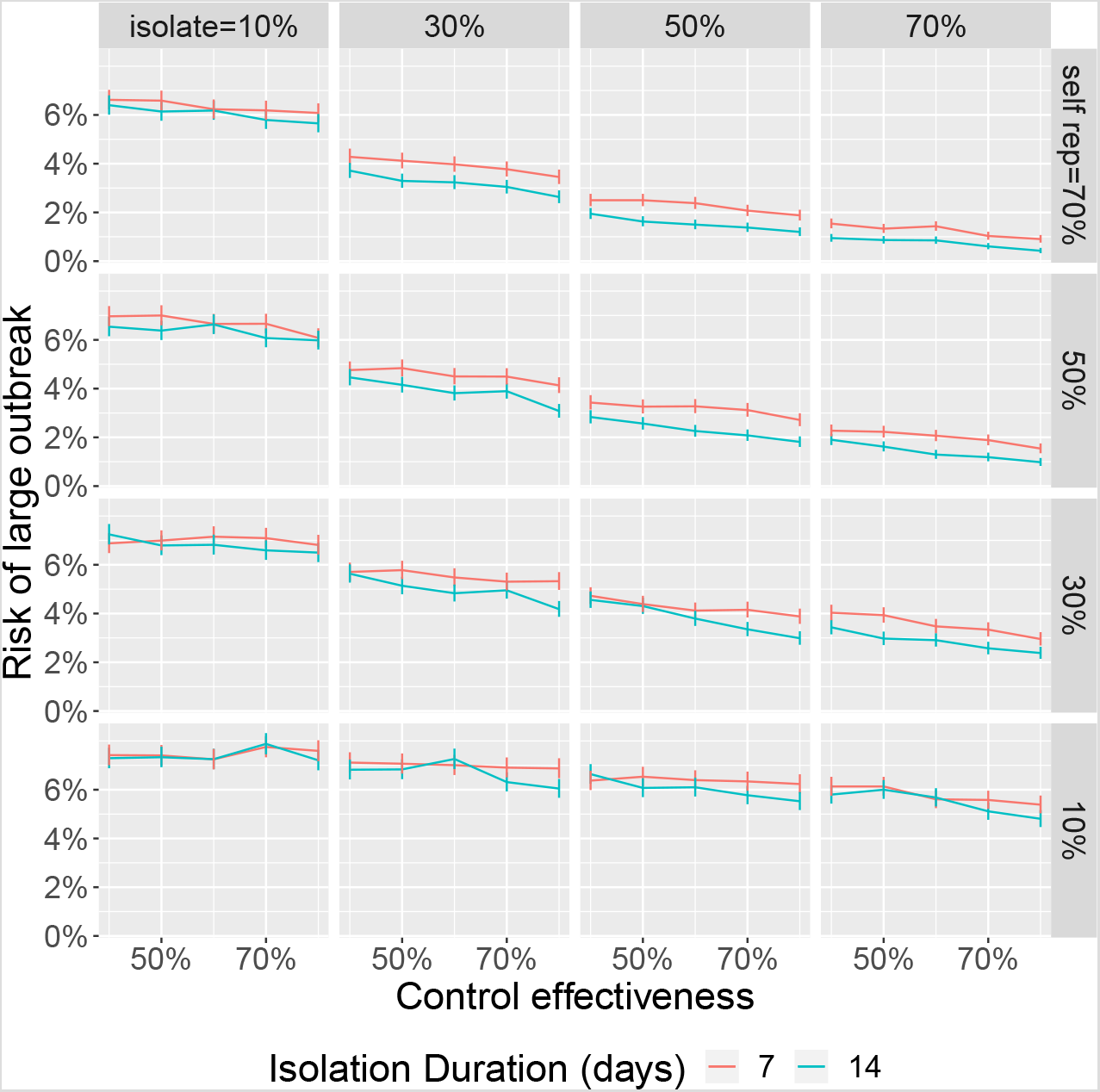
Trade-off between self isolation probability (columns) and self report (rows) with error bars denoting 95% confidence intervals. If we assume we are currently near the top left we expect that introducing legal ramifications for breaking self isolation to move us down and right. Whether this decreases risk depends on the strength of the trade-off.

**Figure S5:**
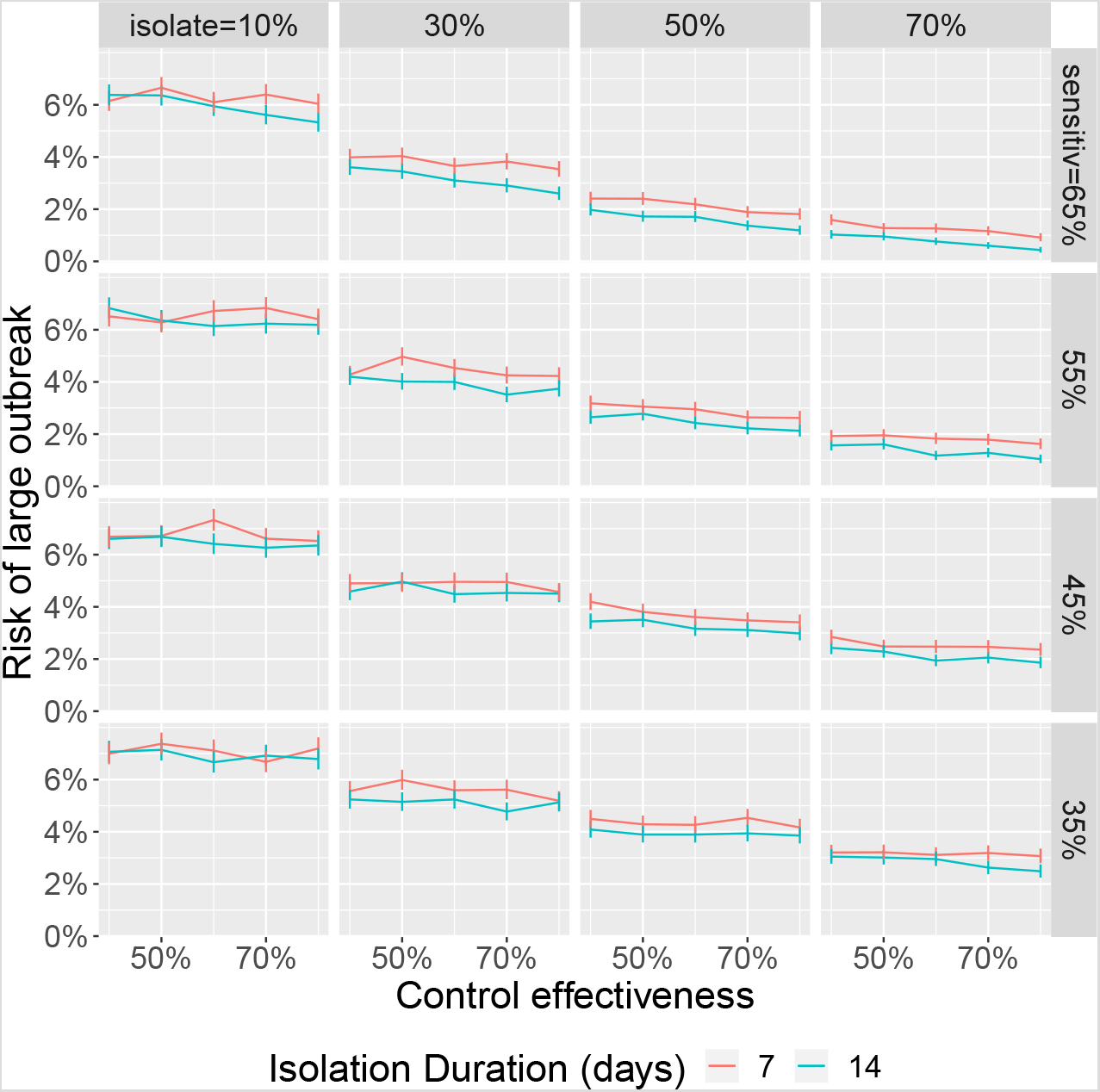
Trade-off between self isolation time (columns) and self report rate (rows) with error bars denoting 95% confidence intervals. Individuals self isolate for a randomly selected duration between min isolation4 and 14 days. Untraced, symptomatic individuals self-report with a probability that varies across the rows. The proportion of close contacts that are divulged and asked to self-isolate varies across the x-axis of each subplot. Self isolation probability is fixed at 70%. If we assume we are currently near the top left we expect that introducing legal ramifications for breaking self isolation to move us down and right. This generally increases risk.

